# Genotype-Based Severity Scoring System in Wolfram Syndrome: Correlation with Onset of Cardinal Symptoms and *WFS1* Gene Variant Types

**DOI:** 10.64898/2026.03.24.26349216

**Authors:** Liam Oiknine, Abby F. Tang, Evan Lee, Nila Palaniappan, Megha Verma, Kasra Vand, Sahiti Somalraju, Sarath Chandra Janga, Fumihiko Urano

## Abstract

Wolfram syndrome is a rare genetic disorder characterized by antibody-negative early-onset atypical diabetes mellitus, optic nerve atrophy, sensorineural hearing loss, diabetes insipidus (arginine vasopressin deficiency), and progressive neurodegeneration, with significant variability in disease severity. We assessed the accuracy of a genotype-based severity scoring system to predict the onset of cardinal symptoms in Wolfram syndrome. This system is based on the type of *WFS1* variants (in-frame or out-of-frame) and their location relative to transmembrane domains. Severity scores were assigned to 324 patients with documented onset ages for diabetes mellitus, optic atrophy, hearing loss, and central diabetes insipidus (arginine-vasopressin deficiency). Our analysis revealed a clear association between the proposed genetic severity scoring system and earlier onset of diabetes mellitus and optic atrophy. Patients with in-frame variants outside transmembrane domains exhibited milder symptoms, especially *WFS1* c.1672C>T (p.Arg558Cys) variant, whereas those with out-of-frame variants showed the earliest onset. Severity scores 3 and 4 did not follow the expected progression, suggesting that transmembrane domain involvement in both alleles may result in greater severity. To independently evaluate the proposed model, we developed a computational framework employing the classification rubric, which was able to confirm that the six-class system can be automatically annotated with 94.4% accuracy. Closer examination of the 7 cases that disagreed with the manual annotation helped improve the annotations and our scoring system, to deploy a three-class model, suggesting the value added by our automatic classifier. Taken together, these findings indicate that the genotype-based severity score is most informative for diabetes mellitus, more modestly informative for optic atrophy, and not currently useful for predicting the onset of hearing loss or diabetes insipidus. Because the analysis is based on observed events without time-to-event censoring and the three-tier consolidation is a post hoc summary of the same dataset, the results should be regarded as exploratory and hypothesis-generating rather than as a fully validated clinical prediction tool, while still offering useful insight into the genotype-related progression of Wolfram syndrome to guide future study.

## 1. Introduction

Wolfram syndrome is a rare autosomal recessive disorder characterized by early-onset antibody-negative diabetes mellitus, optic nerve atrophy, sensorineural hearing loss, diabetes insipidus (arginine vasopressin deficiency) and progressive neurodegeneration [1; 2]. Most patients carry biallelic pathogenic variants in the *WFS1* gene, whereas a smaller subset carry biallelic pathogenic variants in the *CISD2* gene [3]. The objective of this study is to assess the accuracy of our current categorization system for the severity of Wolfram syndrome [4]. Severity is defined by the age of onset of the core manifestations of Wolfram syndrome, with particular focus on diabetes mellitus, which appears to be a key indicator for overall disease severity [4]. The system is based on two features demonstrated to significantly impact the severity of disease manifestations. First, the type of pathogenic variants in the *WFS1* gene has been shown to influence the severity of Wolfram syndrome. Specifically, patients with in-frame missense or insertion/deletion pathogenic variants tend to exhibit milder manifestations than those with frameshift/nonsense pathogenic variants, as evidenced by later onset ages for diabetes mellitus and optic atrophy. Secondly, in-frame variants located within transmembrane domains result in more severe manifestations compared to those outside transmembrane domains.

*WFS1* encodes wolframin, a multi-pass transmembrane glycoprotein anchored in the endoplasmic reticulum (ER) membrane through nine transmembrane segments [5]. Disruption of these segments is expected to compromise membrane insertion, topology, and protein stability, all of which contribute to ER stress and to the pancreatic β-cell, sensorineural, and neuronal vulnerability that characterizes Wolfram syndrome [5; 6; 7; 8]. For this reason, the present scoring framework uses transmembrane-domain location as its second axis: the transmembrane region is the segment for which prior pathogenic-variant clustering and structural data have been jointly reported, not the only functionally important region of the protein. We do not regard the N-terminal cytosolic region or the C-terminal ER-luminal region as unimportant—both clearly contribute to wolframin function—nor do we claim that any single transmembrane segment or connecting loop is more critical than another; the per-segment patient counts in the present cohort are too small to support such claims, and resolving these differences is a goal for a larger, prospectively recruited cohort and for the AlphaFold-based structural modeling described in the Discussion [9; 10]. Finally, because classic Wolfram syndrome is autosomal recessive, this framework is deliberately calibrated for biallelic disease. A spectrum of dominantly inherited *WFS1*-related disorders also exists, ranging from isolated low-frequency sensorineural hearing loss [11; 12], isolated optic neuropathy [13], and isolated cataracts [14] to combinations of hearing loss and optic neuropathy [15; 16; 17] and Hattersley-Urano syndrome characterized by diabetes mellitus, optic atrophy, cataracts, hypotonia, and developmental delay [18]. These autosomal-dominant disorders are related to but clinically distinct from autosomal-recessive Wolfram syndrome, and heterozygous carriers were not included in the present analysis, which is structured around biallelic disease.

## 2. Materials and Methods

### Patients

Subjects, and their parents or legal guardians, as appropriate, provided written, informed consent before participating in this study, which was approved by the Human Research Protection Office at Washington University School of Medicine in St. Louis, MO. Patient data from the Washington University International Registry and Clinical Study for Wolfram Syndrome and patient case reports were analyzed to select for patients with two recessive pathogenic variants in the *WFS1* gene. Patients were excluded if they lacked genetic information for either of their *WFS1* allele variants. Additionally, records were excluded if they did not have a numerical age of onset for their respective clinical phenotype (diabetes insipidus, optic atrophy, diabetes mellitus, hearing loss). Pathogenic variants were then classified as being either nonsense/frameshift variants or missense/in-frame insertion and deletion variants.

Throughout this manuscript, the term “out-of-frame” is used as shorthand for the truncating side of the scoring framework and comprises both frameshift and nonsense (stop-gain) variants. We recognize that nonsense variants are not literally “out-of-frame” in the same sense as frameshift variants; both classes are nevertheless grouped together because both are expected to abolish full-length wolframin expression, through nonsense-mediated decay or premature truncation. We use “truncating” as the equivalent term in the Discussion, and we retain the historical “in-frame/out-of-frame” labels in the figures and tables for continuity with the existing scoring concept [4]. The scoring framework operates one layer downstream of variant classification rather than as a substitute for it: only variants meeting ACMG/AMP criteria for “pathogenic” or “likely pathogenic” were retained, and a variant of uncertain significance (VUS) was included only when the variant on the other allele was pathogenic or likely pathogenic and the patient’s clinical features were consistent with Wolfram syndrome [19]. Synonymous variants were excluded unless they had functionally validated splice consequences; canonical splice-site variants whose predicted effect was loss of the canonical donor or acceptor with frameshift or exon skipping were treated as truncating (out-of-frame side); and deep intronic and non-canonical splice variants were not included because their consequence at the protein level could not be confirmed in our dataset.

### Data Processing and Statistical Analysis

The data used for this analysis was gathered from a sample of more than 400 Wolfram patients, with varying amounts of clinical data. For each patient, the dataset included onset information for canonical Wolfram syndrome symptoms, including diabetes mellitus, optic atrophy, diabetes insipidus, and hearing loss, as well as genetic features such as variant type, allele-specific variant location, and whether the variant is located within a transmembrane domain of the protein. In total, onset information was present for 324, 306, 195, and 149 patients with diabetes mellitus, optic atrophy, hearing loss, and diabetes insipidus, respectively. Patients were divided into subgroups based on the symptoms they had onset ages listed for, and each group was graphed independently excluding outliers. Statistical significance between groups was analyzed using the Kruskal-Wallis test and Dunn’s test with Bonferroni correction. Statistical significance was defined by a p-value < 0.05.

Outliers were defined per phenotype using the Tukey 1.5×IQR rule applied to the within-phenotype distribution of onset ages; the difference between the per-phenotype sample sizes reported above (324, 306, 195, and 149 for diabetes mellitus, optic atrophy, hearing loss, and diabetes insipidus) and the smaller figure-panel sample sizes corresponds exactly to the points removed by this rule. To confirm that outlier removal did not drive our conclusions, we repeated all between-group comparisons with outliers retained as a sensitivity analysis: the direction of every Mild–Moderate–Severe contrast and the rank order of the six severity scores were preserved, and statistical significance was retained for the diabetes mellitus contrasts. Beyond the Kruskal–Wallis and Dunn tests, effect sizes are summarized as differences in median age of onset between groups (reported in the Results) together with the Kruskal–Wallis η² and pairwise Cliff’s δ for the consolidated three-tier comparisons. We did not perform a formal time-to-event (survival) analysis in the present study: standardized last-follow-up dates were not uniformly available in this historical registry and case-report dataset, and phenotype-specific surveillance varied across contributing sites, so right-censoring of patients who had not yet developed a manifestation could not be performed reliably. A formal time-to-event framework—Kaplan–Meier estimation with right-censoring of unaffected patients, Cox proportional-hazards modeling, and an ordinal trend test—is therefore named as the primary planned follow-up analysis once standardized last-follow-up dates are recorded prospectively across the full registry.

### Computational Framework for Rule-Based Severity Score Reconstruction

After the initial genotype–phenotype analysis to manually annotate patients into genotype severity levels, we developed a rule-based computational algorithmic implementation to assign each patient a genotype-based severity score using the six-level *WFS1* framework highlighted in Figure 1. The algorithm first classified each *WFS1* allele separately. When possible, coding DNA insertions, deletions, or duplications were classified by whether the nucleotide change preserved the reading frame. For complex or ambiguous variants, the HGVS protein annotation was used to determine whether the allele produced a frameshift, premature stop codon, missense change, or in-frame alteration [20]. If neither DNA nor protein annotation was sufficient, the recorded variant-class label in the dataset was used as a fallback. Frameshift and nonsense variants were classified as high-severity alleles, whereas missense and in-frame variants were classified as lower-severity alleles. The two allele classifications were then combined with transmembrane-domain status to assign each patient to one of the six genotype severity groups.

**Figure 1.**
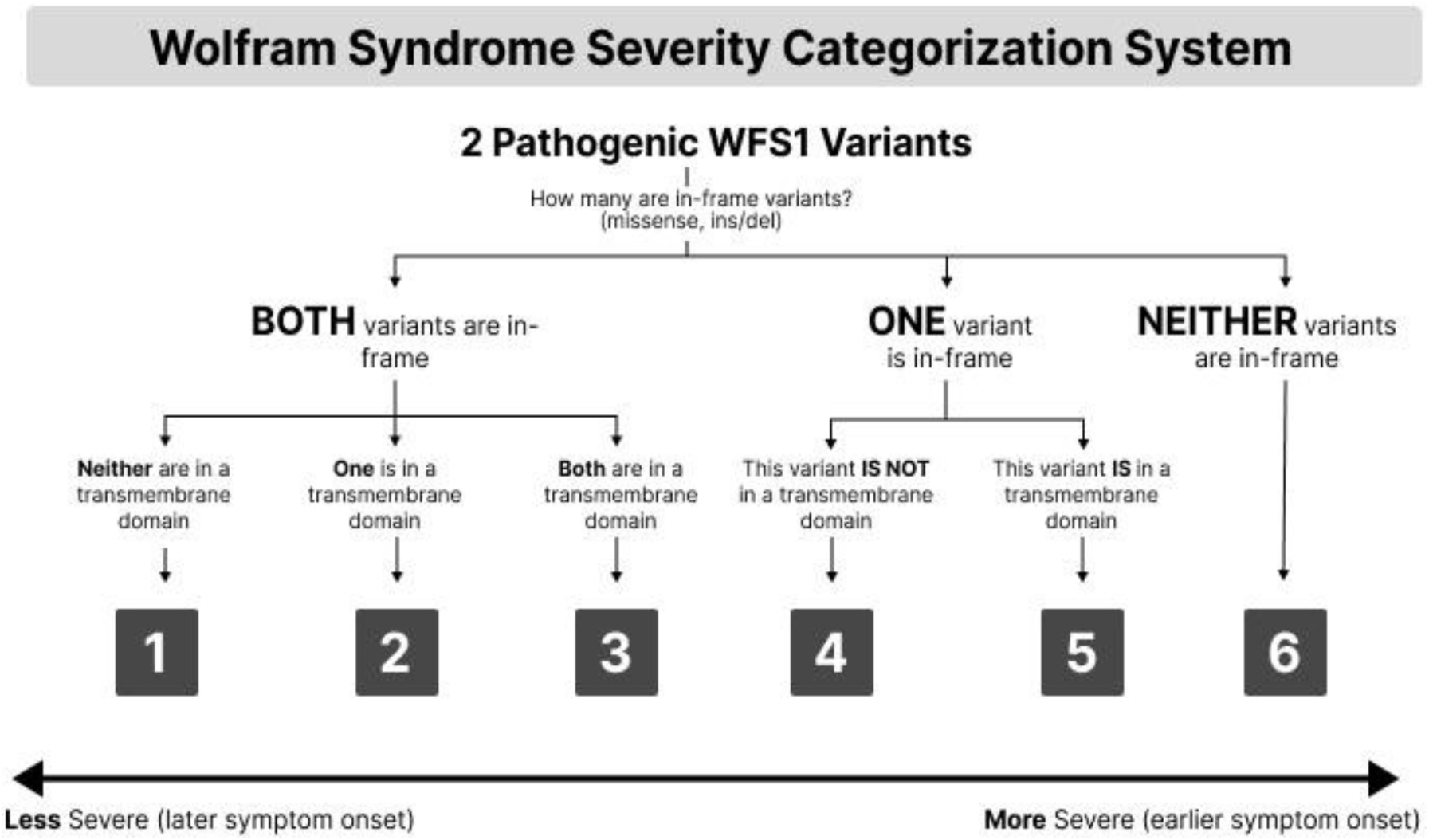
Severity scoring system schematic illustrating the six genotype-based groups ordered by increasing disease severity.

### Machine-learning Validation of Genotype-based Severity Scoring

To evaluate whether the genotype-derived severity score captured the age-at-diagnosis information present in the underlying *WFS1* variants, we performed a genotype-only machine-learning benchmark for diabetes mellitus and optic atrophy. Analyses were restricted to Wolfram syndrome patients with exact recorded diagnosis ages for the phenotype being modeled The diabetes mellitus cohort contained 109 patients with exact age-at-diagnosis values, while the optic atrophy cohort contained 105 patients with exact age-at-diagnosis values.. Clinical phenotype timing other than the target diagnosis age, such as clinical severity labels, visual acuity, C-peptide, and other downstream clinical measurements, were excluded from the predictor set to avoid over-fitting on the relatively small sample size.

Three genotype representations were compared using the same modeling and evaluation framework: the human-annotated genetic severity score alone, the computationally reconstructed 1 to 6 severity score alone, and a broader engineered genotype feature set. The engineered genotype model included mutation-class labels, sorted mutation-pair summaries, amino-acid position features, transmembrane-domain indicators, allele-level frame/consequence summaries, and zygosity or allele-completeness summaries. Categorical variables were imputed and one-hot encoded, and numeric variables were median-imputed and standardized within each training fold (Table 1).

**Table 1.**
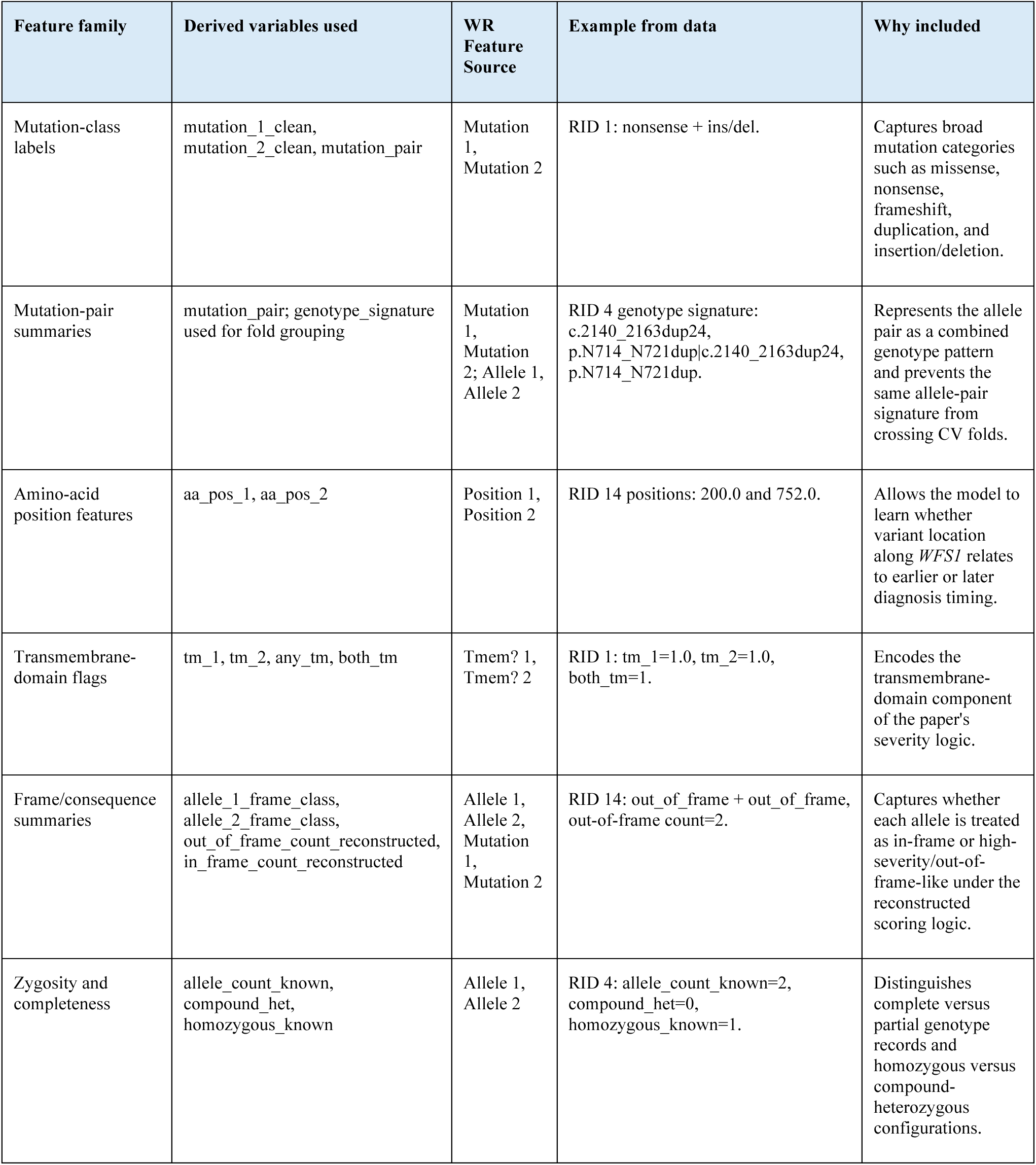
Feature families used in the engineered genotype model.

Gradient boosting regression models were trained separately for diabetes mellitus and optic atrophy diagnosis age. The gradient boosting model used an ensemble of shallow regression trees fit sequentially, so that each additional tree focused on residual prediction error from the previous trees. This allows the model to capture non-linear genotype patterns and interactions, such as combinations of mutation class, transmembrane involvement, and allele position, while remaining restricted to genotype-derived inputs.

Model performance was evaluated using grouped cross-validation, in which patients were grouped by normalized genotype signatures so that individuals sharing the same allele pair were assigned to the same fold. This was done to minimize information leakage arising from shared genotypes between training and testing data. For each held-out patient, the model predicted the exact diagnosis age.

Predicted and observed ages were subsequently categorized into sextile bins using cut points derived exclusively from the training data of each fold. For diabetes mellitus, sextile bins were defined as: ≤4 years, >4-5 years, >5-7 years, >7-9 years, >9-14 years, and >14 years. For optic atrophy, sextile binds were defined as: ≤6.61 years, >6.61-9 years, >9-13 years, >13-15.04 years, >15.04-28 years, and >28 years. Performance was summarized as exact sextile accuracy, within-one-sextile accuracy, and mean absolute sextile error.

## 3. Results

### Genotype severity framework for categorizing Wolfram Syndrome

When the type of *WFS1* variants and their location relative to transmembrane domains are considered together [21], they form six distinct groups of patients, ordered by increasing disease severity. The severity scores are defined as follows (**Figure 1**)( https://severity-scoring.vercel.app/calculator):

- **Severity Score 1:** Both alleles contain in-frame (missense, ins/del) variants; neither variant lies within a transmembrane domain.
- **Severity Score 2:** Both alleles contain in-frame (missense, ins/del) variants; one variant lies within a transmembrane domain, the other does not.
- **Severity Score 3:** Both alleles contain in-frame (missense, ins/del) variants; both variants lie within transmembrane domains.
- **Severity Score 4:** One allele contains an in-frame (missense, ins/del) variant, the other contains an out-of-frame (frameshift/nonsense) variant. The in-frame variant is not located in a transmembrane domain.
- **Severity Score 5:** One allele contains an in-frame (missense, ins/del) variant, the other contains an out-of-frame (frameshift/nonsense) variant. The in-frame variant is located in a transmembrane domain.
- **Severity Score 6:** Both alleles contain out-of-frame (frameshift/nonsense) variants.

Using the severity scoring system, we evaluated the association between disease severity and age of onset. Diabetes mellitus followed a clear trend with earlier ages of onset correlating to higher severity scores, the only exception being severity score 3. Statistical significance was seen between scores 2 and 3 (p = 0.008128) as well as scores 4 and 5 (p = 0.001535), serving as strong breaking points for consolidating the data into three groups (**Figure 2, upper panel, Table 2**, **Table 3**). The ‘Mild’ group consists of severity scores 1–2, the ‘Moderate’ group consists of severity scores 3–4, and the ‘Severe’ group consists of severity scores 5–6. Consolidating these groups allowed for statistical significance to be achieved between each of the three groups (Mild vs. Moderate: p = 0.011445; Moderate vs. Severe: p = 5.25 × 10^−6^). These groups, with median onset ages of 9.0, 6.5, and 4.6 years for ‘Mild,’ ‘Moderate,’ and ‘Severe’ respectively, may serve as accurate benchmarks for predicting DM onset ages based on genotype (**Figure 2, lower panel**). <u>Consolidation of severity scores 1–6 into the Mild (1 and 2), Moderate (3 and 4), and Severe (5 and 6) tiers was guided by the same dataset that revealed the significant adjacent contrasts; this three-tier grouping should therefore be regarded as a data-driven, post hoc summary rather than an independently validated classification, and the performance of the three-tier model is likely to represent an upper bound until it is tested in an independent cohort.</u> Optic atrophy follows a somewhat linear trend, though less consistently than diabetes mellitus. Group 3 again shows an earlier onset compared to group 4. Near statistical significance was observed between adjacent groups 3 and 4 (p = 0.059694), but no two adjacent groups reached significance (**Figure 3, upper panel, Table 2**, **Table 3**). The data was consolidated into three subgroups (‘Mild’, ‘Moderate’, and ‘Severe’) according to the same breakdown of severity scores as defined above, with median onset ages of 14.0, 11.0, and 9.0 years respectively (**Figure 3, lower panel**). This representation shows a clearer trendline correlating higher severity scores to earlier onset of optic atrophy, though these groups are less clearly defined compared to those of diabetes mellitus. A potential contributing factor is that optic atrophy may only be noticed gradually after onset, potentially introducing reporting bias in the recorded onset ages. Hearing loss and diabetes insipidus data do not follow any specific pattern, indicating no correlation between age of onset and the current severity scoring system. This is consistent with prior studies showing that neither variant type nor transmembrane domain location significantly influences these manifestations. Notably, both hearing loss and diabetes insipidus graphs share similar shapes, with earlier onset ages observed in patients with scores 2 and 5 (**Figure 5**, **Table 2**, **Table 3**).

**Figure 2.**
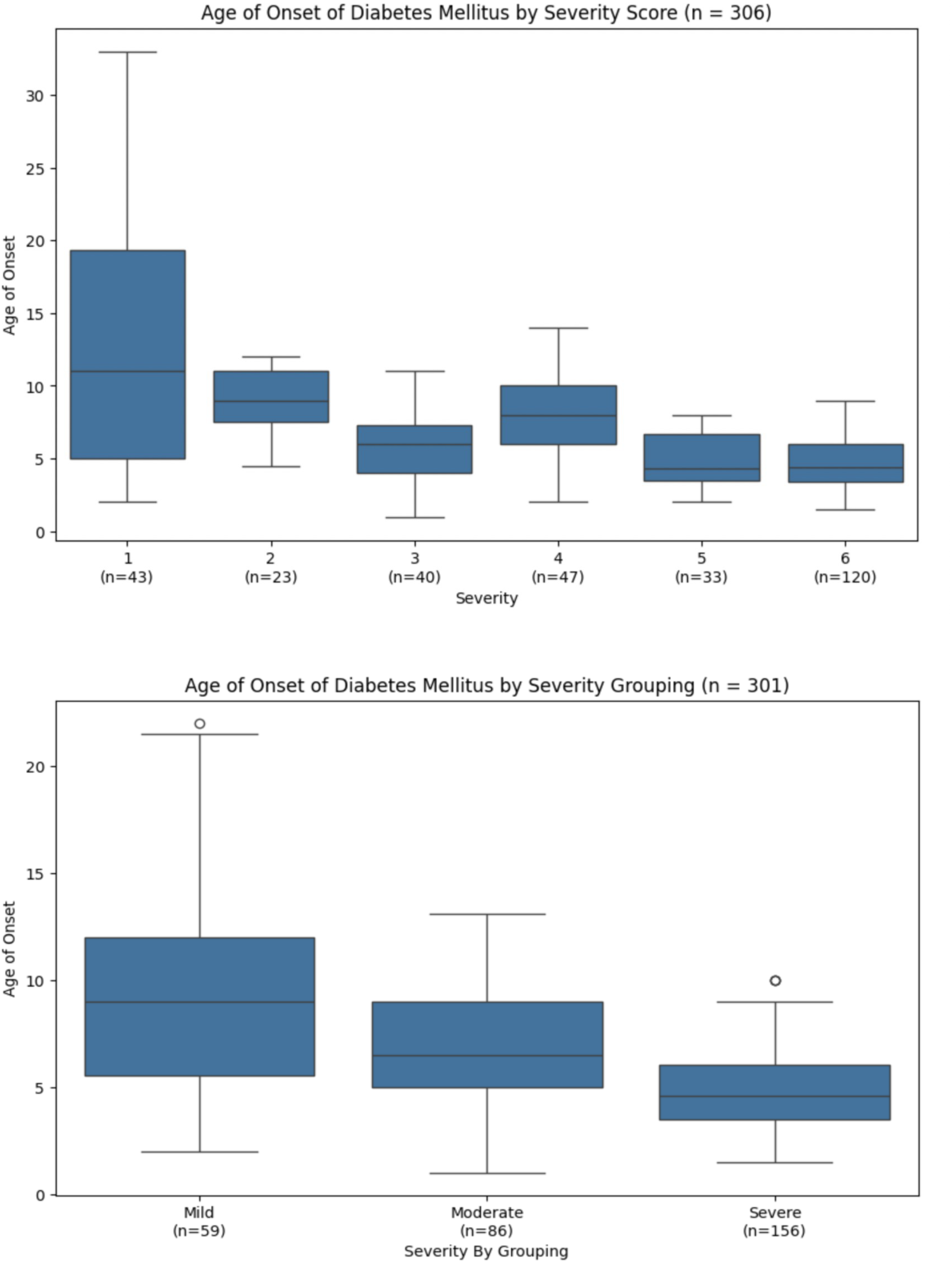
Diabetes Mellitus onset age by individual severity score (top) and by consolidated Mild/Moderate/Severe groups (bottom). Statistical significance between scores 2 and 3 (p = 0.008128) and scores 4 and 5 (p = 0.001535). Consolidated group comparisons: Mild vs. Moderate p = 0.011445; Moderate vs. Severe p = 5.25 × 10^−6^. Median onset ages: 9.0, 6.5, and 4.6 years for ‘Mild’, ‘Moderate’, and ‘Severe’ respectively.

**Figure 3.**
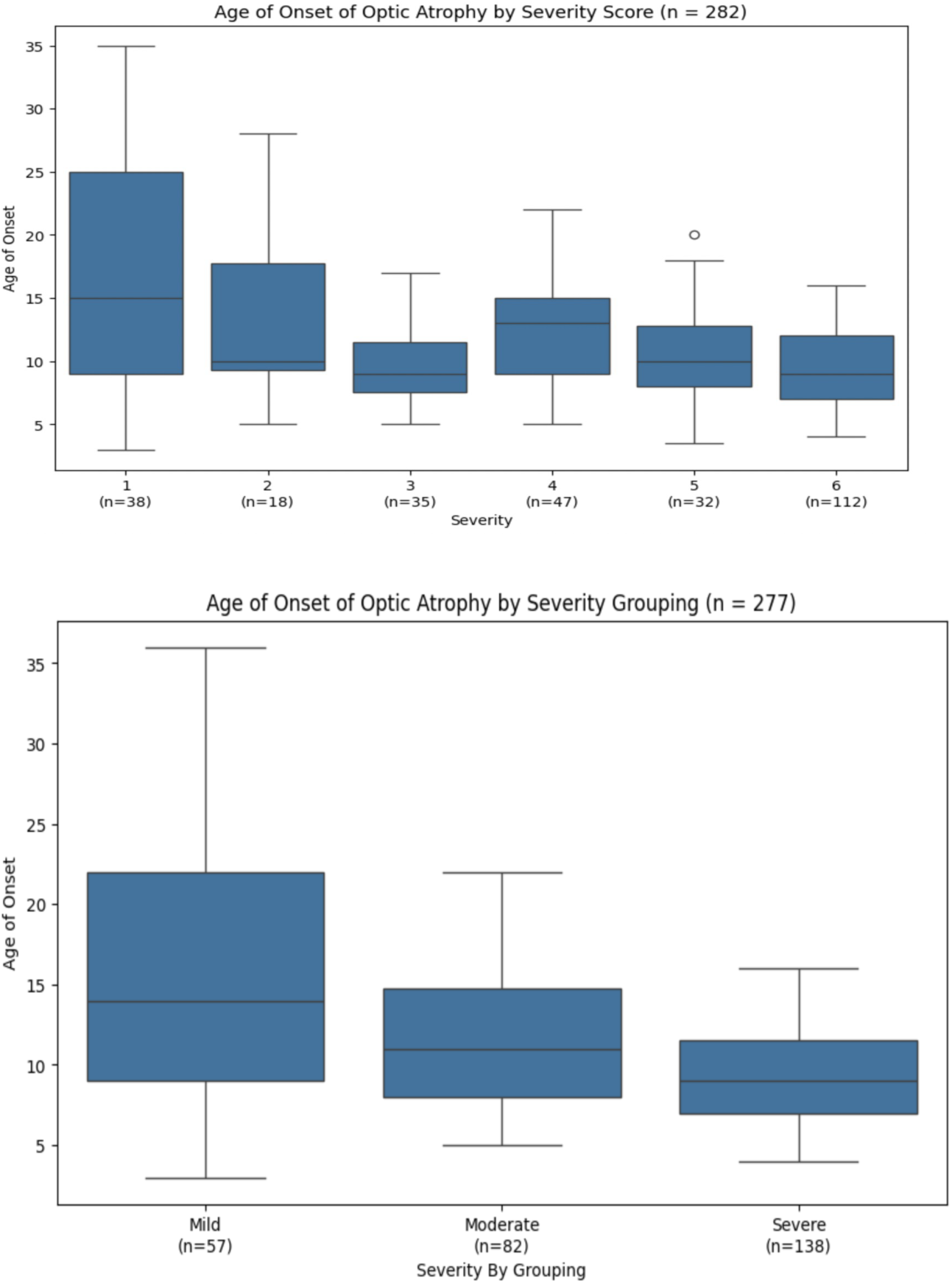
Optic Atrophy onset age by individual severity score (top) and by consolidated Mild/Moderate/Severe groups (bottom). Near statistical significance between groups 3 and 4 (p = 0.059694). Median onset ages: 14.0, 11.0, and 9.0 years for ‘Mild’, ‘Moderate’, and ‘Severe’ respectively.

**Table 2.** Median Age of Onset – Full Dataset.

|  | **Diabetes Mellitus** | **Optic Atrophy** | **Hearing Loss** | **Diabetes Insipidus** |
| --- | --- | --- | --- | --- |
| Full Dataset (n) | 324 | 306 | 195 | 149 |
| Median Onset Age (years) | 6.0 | 11.0 | 14.0 | 13.0 |

**Table 3.** Median and Mean Age of Onset by Severity Score. DM = Diabetes Mellitus; OA = Optic Atrophy; HL = Hearing Loss; DI = Diabetes Insipidus

| Severity Score | Median Age of Onset (years) |  |  |  | Mean Age of Onset (years) |  |  |  |
| --- | --- | --- | --- | --- | --- | --- | --- | --- |
|  | DM | OA | HL | DI | DM | OA | HL | DI |
| 1 | 11.0 | 15.0 | 15.5 | 14.0 | 13.0 | 16.9 | 16.4 | 13.3 |
| 2 | 9.0 | 10.0 | 8.0 | 11.5 | 9.0 | 13.2 | 8.0 | 11.5 |
| 3 | 6.0 | 9.0 | 11.0 | 14.6 | 5.9 | 9.8 | 11.1 | 16.4 |
| 4 | 8.0 | 13.0 | 15.0 | 12.0 | 7.8 | 12.7 | 15.5 | 12.6 |
| 5 | 4.3 | 10.0 | 10.0 | 9.8 | 4.8 | 10.6 | 11.7 | 13.0 |
| 6 | 4.4 | 9.0 | 13.0 | 12.0 | 4.9 | 9.5 | 14.3 | 12.4 |

To further evaluate our proposed genetic severity scoring system, we have applied a computational rule-based reconstruction algorithm to the final diabetes mellitus/optic atrophy timing cohort (see Materials and Methods), which exhibited a high agreement with annotated scores. Among 125 Wolfram syndrome patients with both scores available, 118 retained the same six-level score (94.4%), and 119 remained in the same mild/moderate/severe group (95.2%). Seven patients changed score after reconstruction (RID 23, 143, 243, 265, 266, 270, and 313), most commonly due to discrepancies between the coding DNA annotation, HGVS protein consequence, and human annotated mutation-class labels (**Table 4**).

**Table 4.**
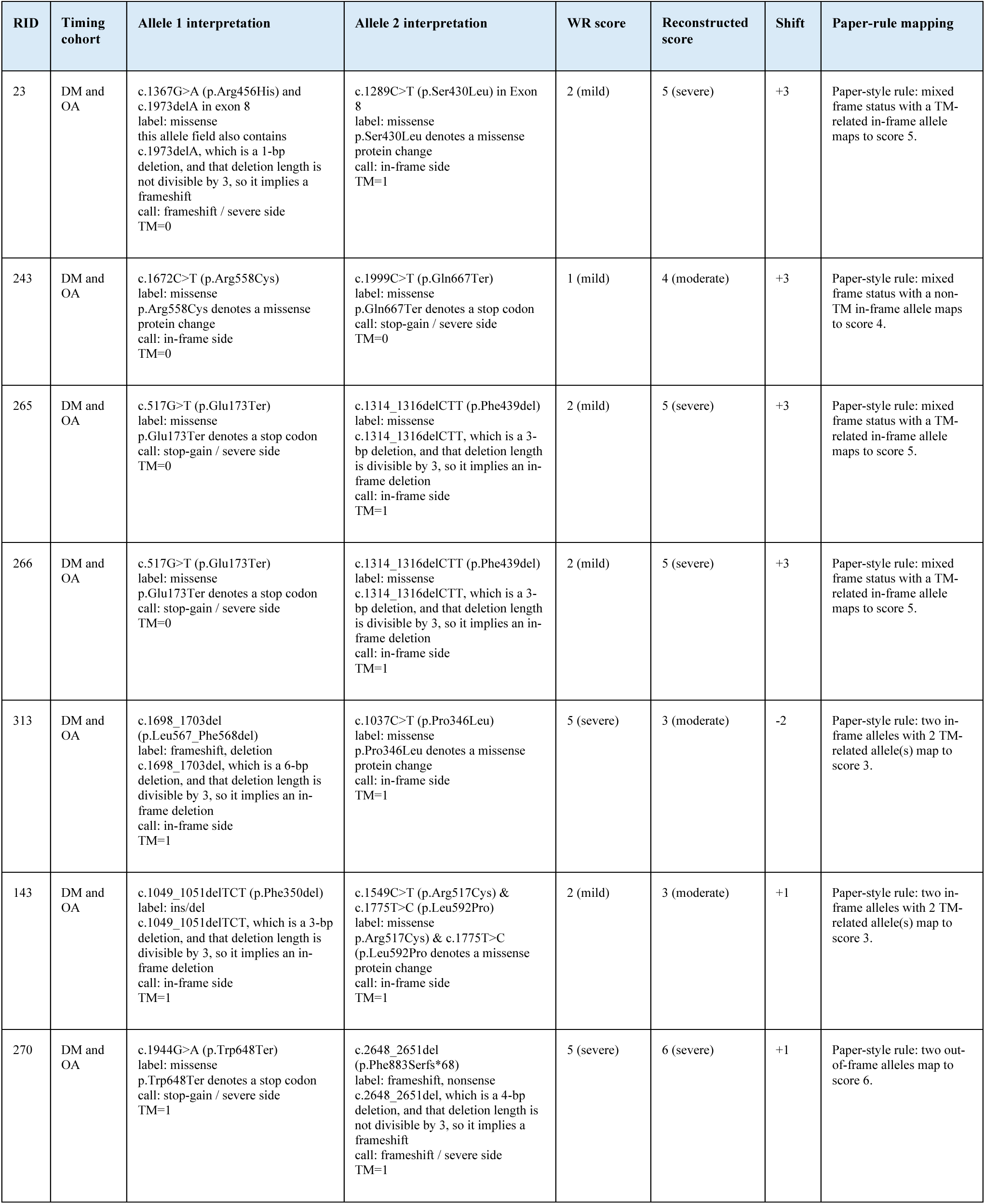
Final-cohort patients with discordant WR provided and reconstructed severity scores. Each row lists one Wolfram syndrome patient from the final diabetes mellitus/optic atrophy timing cohort whose WR provided severity score differed from the rule-based reconstructed score. Allele interpretations show the annotation used for reconstruction, the WR mutation label, the inferred consequence side, and transmembrane-domain status. *DM, diabetes mellitus; OA, optic atrophy; RID, registry identifier; TM, transmembrane domain*.

### Machine learning model based on gradient boosting exhibited the highest scoring accuracy for predicting the age of onset

To further evaluate whether genotype-derived severity information could predict clinical timing, genotype-only machine-learning models were developed to estimate age of onset for diabetes mellitus and optic atrophy (see Materials and Methods). The objective of these models was to determine how well genetic severity representations alone could capture age-of-diagnosis patterns across Wolfram syndrome patients, while excluding downstream clinical measurements to avoid information leakage.

For validating the genotype-only machine-learning, the full engineered genotype model was compared with the original Wolfram syndrome and related disorders registry (WR) score and the reconstructed score. For diabetes mellitus, the engineered genotype model improved sextile placement compared with the original score from WR cohort, increasing exact sextile accuracy from 19.3% to 29.4% and within-one-sextile accuracy from 60.6% to 69.7% (**Figure 4**, **Table 5**). For optic atrophy, the engineered genotype model also improved performance relative to the score from WR cohort, increasing exact sextile accuracy from 15.2% to 18.1% and within-one-sextile accuracy from 45.7% to 57.1%. These findings suggest that the WR score captures meaningful genotype-timing signal, while richer genotype features retain additional predictive information hinting to strong association between the patients’ genotypic signature and Wolfram syndrome’s phenotypic patterns.

**Figure 4.**
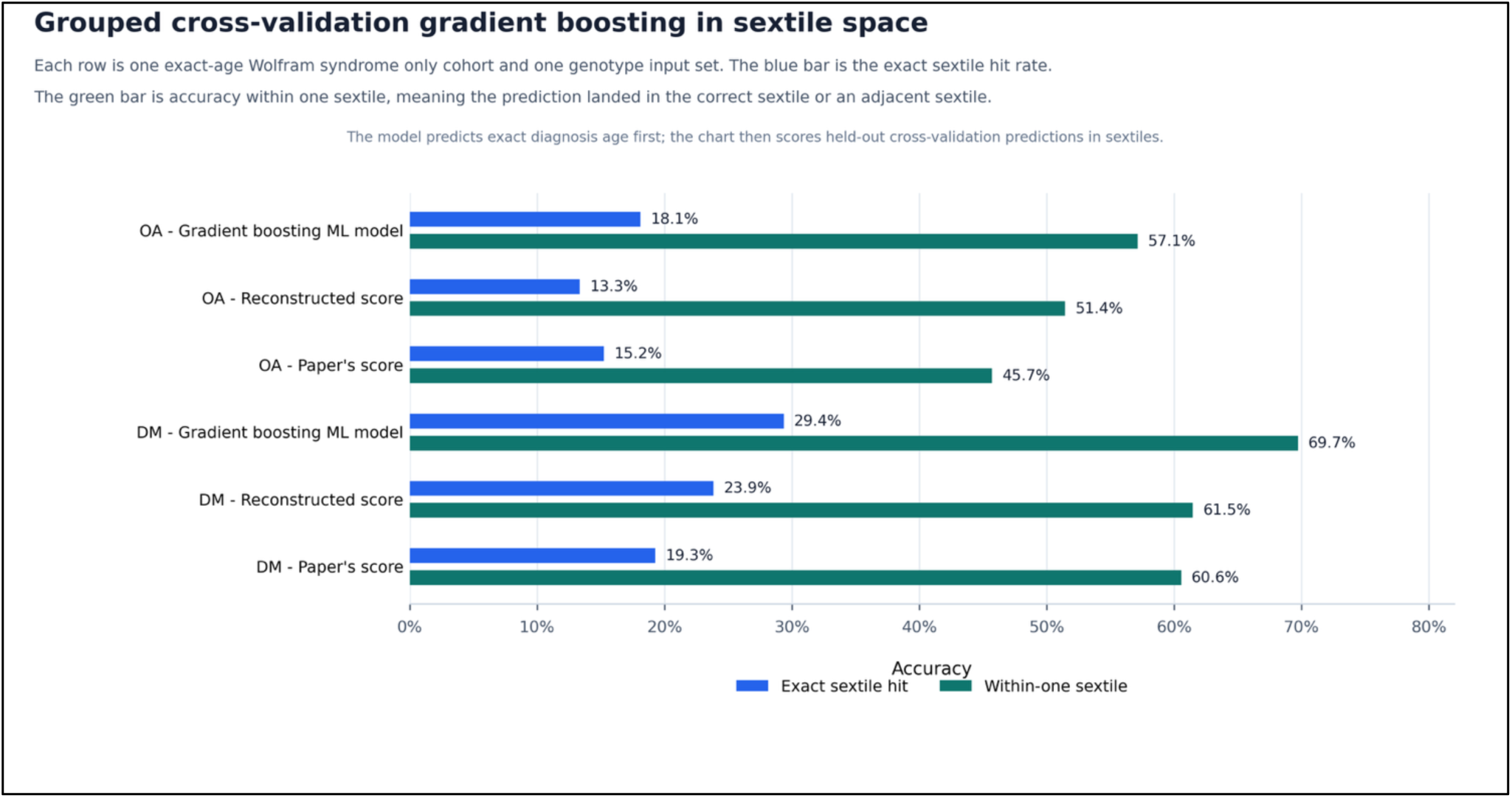
Genotype-only machine-learning validation of severity scoring for diabetes mellitus and optic atrophy. Gradient boosting regression models were trained under grouped cross-validation and evaluated by mapping held-out exact age predictions into sextiles. The WR paper score, reconstructed severity score, and full engineered genotype model were compared using exact sextile accuracy and within-one-sextile accuracy. The full engineered genotype model used mutation-class labels, mutation-pair summaries, amino-acid position summaries, transmembrane-domain flags, frame/consequence summaries, and zygosity or allele-completeness summaries, while excluding clinical phenotype timing, visual acuity, C-peptide, and other downstream clinical measurements.

**Figure 5.**
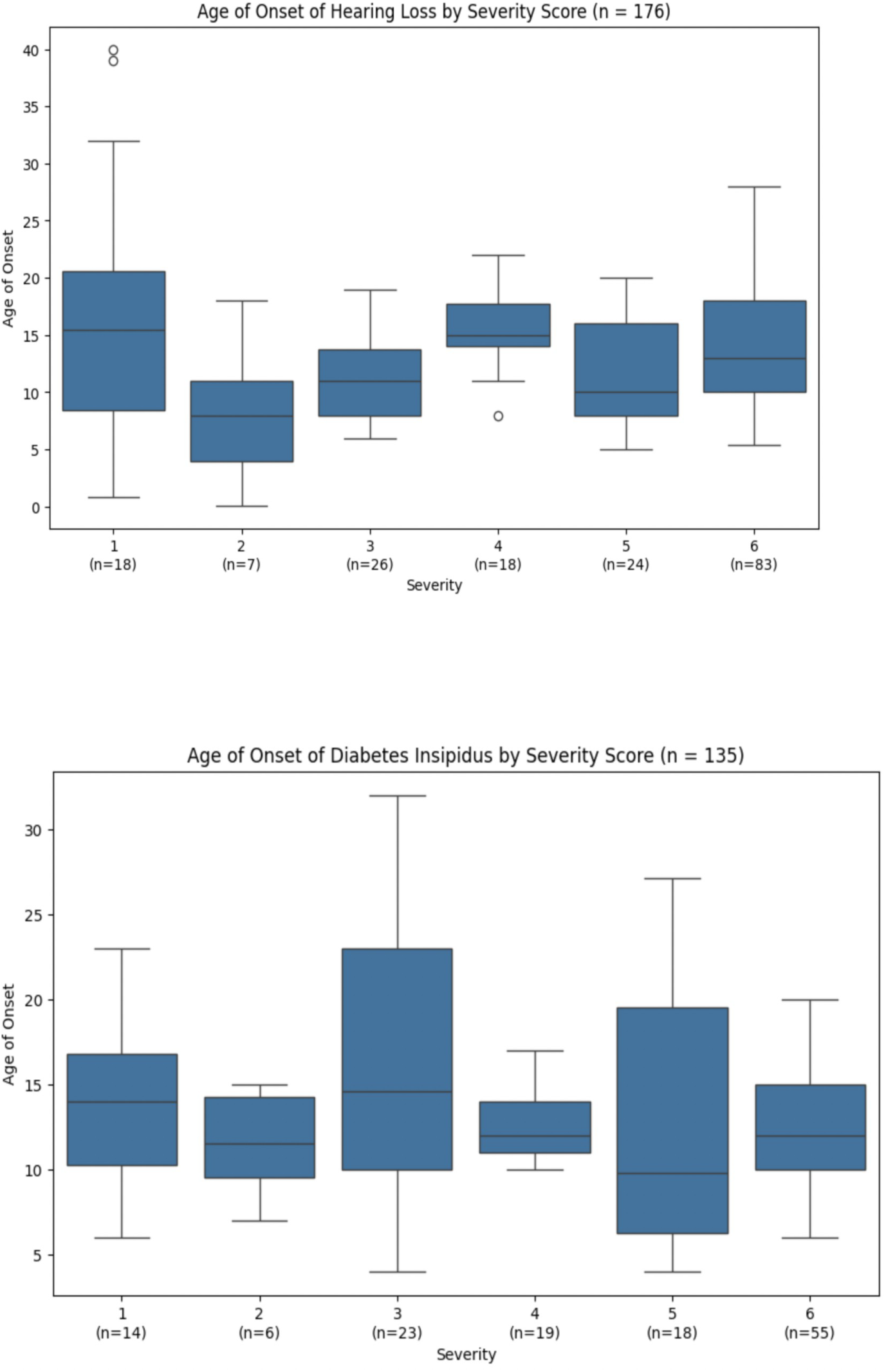
Hearing Loss (top) and Diabetes Insipidus (bottom) onset age by severity score. No statistical significance between any adjacent groups and no consistent pattern is apparent.

**Table 5.**
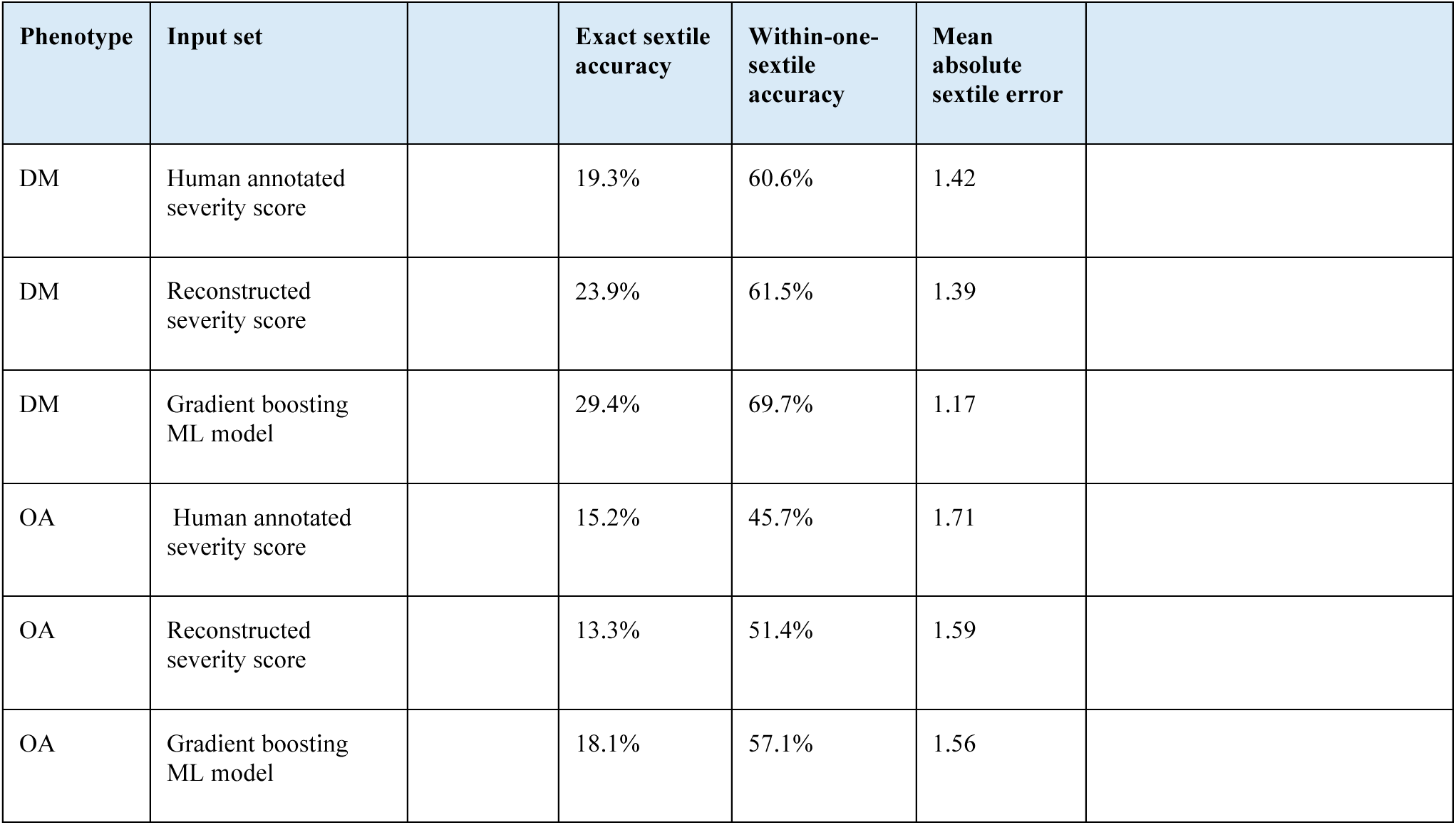
Sextile performance metrics for the genotype-only. **Figure 4 benchmark**

## 4. Discussion

Our scoring system demonstrates consistency for diabetes mellitus and optic atrophy, and shows no significant correlation with age of onset for hearing loss or diabetes insipidus. While the system provides prognostic insight for these two cardinal manifestations, further refinement is needed to improve its predictive accuracy and broaden its clinical utility.

Several limitations temper the interpretation of these findings and should be addressed before the score is applied clinically. First, the present cohort is drawn predominantly from the Washington University International Registry and Clinical Study for Wolfram Syndrome and related disorders and from previously published case reports, and it partially overlaps with the dataset used to derive the original six-level scoring concept [4]. The present study should therefore be read as an internal refinement and confirmation of the scoring framework rather than as a fully independent external validation; a prospective replication in a non-overlapping multinational cohort will be required before the score is recommended for clinical decision-making. For the same reason, the consolidation of the six scores into three tiers, which was guided by this dataset, is best regarded as exploratory.

Second, age of onset is only an indirect proxy for biological severity. It is influenced not only by the molecular consequences of *WFS1* variants but also by ascertainment, screening intensity, clinical vigilance, and the gradual detectability of optic atrophy, sensorineural hearing loss, and diabetes insipidus compared with the more clinically conspicuous onset of diabetes mellitus. The reporting bias we previously noted for optic atrophy, therefore, applies, in attenuated form, to all four endpoints, and absolute median onset ages should be interpreted with this caveat in mind. Future severity scales would benefit from being benchmarked against orthogonal severity readouts—residual C-peptide trajectory [22], neurofilament light chain [23; 24], visual-field and retinal-nerve-fiber-layer measures [25; 26; 27; 28], and audiometric thresholds [29]—rather than relying on a single age-of-onset endpoint per phenotype.

Third, the relationship between the present scoring framework and the ACMG/AMP variant-classification system warrants emphasis [19]. The score operates on top of ACMG/AMP-classified pathogenic and likely pathogenic variants and is not designed to up-classify variants of uncertain significance; a formal table of ACMG/AMP categories for each individual variant in the cohort is beyond the scope of the present manuscript but is a logical next step. Relatedly, although the six levels are presented in an order of approximately increasing severity, the observation that score 3 behaves as more severe than score 4 for diabetes mellitus, and to a lesser extent for optic atrophy, means that the six-level scheme is most appropriately interpreted as a categorical genotype classification with an approximate severity ordering rather than as a strictly monotonic ordinal scale. We therefore recommend that the consolidated three-tier scheme be used when an ordinal interpretation is desired; within that scheme, scores 3 and 4 both fall in the Moderate tier, so the practical impact of the non-monotonicity is small but real.

Taken together, these limitations argue for a tempered interpretation of the present results. The score is best understood as a research instrument that is most informative for diabetes mellitus, more modestly informative for optic atrophy, and not currently useful for predicting hearing loss or diabetes insipidus. The forward-looking applications described below—prognostic counseling, trial enrollment, and personalized medicine—are aspirations for future iterations that will incorporate time-to-event analyses, external validation cohorts, ACMG-compatible variant calls, functional ER-stress assays, and additional biomarker readouts, rather than established capabilities of the current version. Accordingly, the findings reported here should be regarded as exploratory and hypothesis-generating.

Several avenues may guide this refinement. Wolfram syndrome is considered a prototype endoplasmic reticulum (ER) disease [30; 31], and functional assays quantifying ER stress levels associated with individual *WFS1* variants could provide a more granular and biologically grounded basis for severity classification [32]. In addition, circulating biomarkers measured in our patients, including C-peptide and neurofilament light chain, as well as emerging ER stress markers such as mesencephalic astrocyte neurotrophic factor, may help stratify disease severity more precisely and refine score boundaries [24; 33; 34]. Structural modeling of each variant using tools such as AlphaFold may similarly help predict functional impact, including effects on protein stability, membrane insertion, and inter-domain interactions, beyond what variant type and transmembrane domain location alone can capture [9; 10]. Finally, continued patient recruitment to our international registry (https://wolframsyndrome.wustl.edu/), along with systematic collection of genetic and longitudinal clinical data, will expand the sample sizes needed to draw more robust conclusions, particularly for hearing loss and diabetes insipidus, where our current cohort is likely underpowered to detect meaningful genotype-phenotype relationships.

A notable anomaly within the scoring framework is that severity score 3, defined by two in-frame variants both located within transmembrane domains, appears more severe than score 4, which combines one frameshift or nonsense variant with one in-frame variant outside a transmembrane domain. This is counterintuitive given that frameshift and nonsense variants are generally expected to produce greater loss of *WFS1* function than in-frame variants. One possible explanation is that simultaneous disruption of transmembrane domains on both alleles impairs protein folding and membrane topology more severely than a single truncating variant paired with a partially functional allele, perhaps because residual *WFS1* activity from the milder allele in score 4 patients is sufficient to provide a degree of functional compensation. Consistent with this interpretation, severity score 3 is associated with earlier median onset ages for diabetes mellitus, optic atrophy, and hearing loss relative to score 4. Repositioning scores 3 and 4 in the rating system may therefore improve its face validity, though this reordering has limited practical consequences under the consolidated three-tier grouping, in which both scores fall within the ‘Moderate’ category, which performs robustly despite this internal discrepancy.

The reconstruction audit also highlights a practical limitation of applying the scoring system retrospectively: variant annotations are not always internally consistent across coding DNA notation, HGVS protein consequence, and mutation-class labels. A reproducible parsing hierarchy can reduce this ambiguity and make score assignment more transparent. Importantly, only a small fraction of final-cohort patients changed score after reconstruction, and most remained in the same mild/moderate/severe category, supporting the robustness of the consolidated three-tier classification while identifying specific cases for expert adjudication.

Given that hearing loss and diabetes insipidus show no correlation with the current rating scale, it is possible that the genetic and molecular determinants of these manifestations may be distinct from those driving diabetes mellitus and optic atrophy onset. Future work aimed at identifying variant-specific effects on auditory hair cell function and arginine vasopressin-producing neurons may ultimately support the development of separate, manifestation-specific severity scales that better capture the full phenotypic complexity of Wolfram syndrome.

This study provides a systematic evaluation of a genotype-based severity scoring system in Wolfram syndrome, applied to one of the largest patient cohorts examined to date. The results demonstrate that *WFS1* variant type and transmembrane domain involvement are useful predictors of diabetes mellitus and optic atrophy onset, and that a consolidated three-tier severity classification, comprising Mild, Moderate, and Severe groups, potentially offers a clinically practical framework for prognostic counseling. These findings represent a step toward personalized medicine in Wolfram syndrome, where early and accurate prediction of disease trajectory could inform the timing of clinical monitoring, guide enrollment in interventional trials, and support family counseling at the time of diagnosis. As therapeutic strategies targeting ER stress and *WFS1* function continue to advance [35; 36; 37], a well-validated severity scoring system will be an increasingly valuable tool for patient stratification and outcome assessment. We anticipate that integration of functional, structural, and biomarker data into future iterations of this system will substantially improve its scope and predictive power across all major manifestations of this devastating disease.

## Author Contributions

Conceptualization: FU and SCJ. Data curation: LO, AT, EL, NP, and MV. Investigation: LO, AT, EL, NP, and MV. Software: KV and SS. Formal analysis: KV, SS, and LO. Methodology: KV, SS, SCJ, and FU. Validation: KV, SS, and SCJ. Visualization: KV and SS. Resources: FU. Funding acquisition: FU. Supervision: FU and SCJ. Project administration: FU and SCJ. Writing – original draft: LO and FU. Writing – review and editing: all authors. All authors read and approved the final version.

## Acknowledgments

We sincerely thank the members of the Washington University Wolfram Syndrome and Related Disorders Clinic and Research Team (https://wolframsyndrome.wustl.edu) for their invaluable support. We are especially grateful to all participants in the Wolfram Syndrome and Related Disorders International Registry, Clinical Study, and Clinical Trials for their time, dedication, and commitment to advancing research. We also gratefully acknowledge the philanthropic support and encouragement from the Auerbach Hyman Fund, the Philipp Fund, Jerome W. Gratenstein Memorial Foundation, the WAVE fund, the WAV fund, the Silberman Fund, the Stowe Fund, the Feiock Fund, the Cachia Fund, the Gildenhorn Fund, the Snow Foundation, the Ellie White Foundation for the Rare Genetic Disorders, the Unravel Wolfram Syndrome Fund, Be A Tiger Foundation, the Eye Hope Foundation, Ontario Wolfram League, Associazione Gentian Sindrome di Wolfram Italia, Alianza de Familias Afectadas por el Sindrome Wolfram Spain, Wolfram Heroes Society, Wolfram syndrome UK, and Association Syndrome de Wolfram France.

## Data Availability Statement

The clinical data supporting the findings of this study are not publicly available due to patient privacy considerations and restrictions of the IRB-approved protocols governing the Washington University International Registry and Clinical Study for Wolfram Syndrome. Deidentified data may be made available from the corresponding author, Fumihiko Urano, MD, PhD, upon reasonable request and subject to institutional review and execution of an appropriate data use agreement.

